# Altered Dynamics and Characterization of Functional Networks in Cocaine Use Disorder: A Coactivation Pattern Analysis of Resting-State fMRI data

**DOI:** 10.1101/2024.06.18.24309063

**Authors:** Benjamin Klugah-Brown, Xing Yao, Hang Yang, Pan Wang, Bharat B. Biswal

## Abstract

**Background:** Cocaine Use Disorder (CUD) poses significant neurobiological and neuropsychiatric challenges, often resulting in severe cognitive and behavioral impairments. This study aims to explore the neural dynamics of CUD using a dynamic coactivation pattern (CAP) analysis approach to provide a deeper understanding of the transient neurobiological mechanisms of the disorder.

**Methods:** Resting-state functional MRI data (SUDMEX_CONN) from 56 CUD patients and 57 healthy controls (HC) were analyzed. CAP analysis was employed to capture transient brain states and their coactivation patterns. Temporal dynamic metrics such as Fraction of Time, Persistence (PST), and Counts were computed to assess differences between groups. Stationary functional connectivity (sFC) was also examined, and meta-analytic term mapping from the Neurosynth database was used to characterize functional associations.

**Results:** CAP analysis revealed six distinct coactivation patterns, with five showing high spatial similarity between CUD and HC groups. Notable differences were observed in State 6, which displayed inverse activation patterns between the groups. CUD individuals exhibited significantly reduced PST across all brain states and altered transition probabilities, particularly increased transitions from the default mode network (DMN) to the somatomotor network and decreased transitions from DMN to attentional/executive networks. Clinical correlations indicated that prolonged cocaine use was associated with altered PST in specific brain states. sFC analysis identified significant alterations in regions such as the right supramarginal gyrus, left superior frontal gyrus, right precentral gyrus, and right lingual gyrus, each linked to distinct cognitive and behavioral functions.

**Conclusions:** This study highlights the utility of CAP analysis in capturing the dynamic neural underpinnings of CUD. The findings provide insights into the neurobiological mechanisms of the disorder, suggesting potential biomarkers for CUD. These results have implications for developing an enhanced approach for substance use disorders, as well as improving our understanding and management of CUD.

## Background

Cocaine addiction as a global threat is particularly problematic because of its profound neurobiological and neuropsychiatric effects. Cocaine use disorder (CUD) is characterized by an intense and compulsive urge to use, often despite knowledge of the negative consequences. This disorder is often accompanied by significant deterioration in cognitive and behavioral health, which exacerbates socioeconomic consequences [1]. Therefore, exploring the neural origins of CUD could lead to better diagnostic and therapeutic strategies not only for cocaine addiction but for other illicit substances.

Advanced neuroimaging studies have demonstrated significant changes in brain activity in different regions in cocaine users [2, 3]. Particularly, resting-state functional magnetic resonance imaging (rs-fMRI) has improved our understanding of brain network dynamics in health and disease [4, 5]. Such imaging techniques have classically revealed functional connections consistent with known structural mappings [6]. A reduction in resting-state functional connectivity is found between several key brain regions, such as the ventral striatum, insula, dorsolateral prefrontal cortex and cerebellum, indicating dysfunction in brain activity and connectivity [7] and highlighting the intricate neural connections in CUD. Moreover, our previous functional and structural meta-analyzes [8, 9] have shown that activation and volumetric alterations in CUD overlap with other addictions.

The traditional stationary functional connectivity (sFC) approach assumes stable cerebral architecture during an fMRI scan, akin to structural connectivity studies [6, 10]. Dynamic FC (dFC), in contrast, reveals transient brain States crucial for understanding cocaine responses. Neuronal dynamic connectivity properties emphasize its context-dependent evolution, balancing efficient processing with metabolic demands [11]. These dynamics, intertwined with cognition and emotion, offer deeper insights into cocaine addiction. Examining connectivity with dFC provides neurobiological perspectives, particularly in studying mechanisms underlying cocaine-addicted brains.

Recent methodological advances in dFC include a dynamic coactivation pattern (CAP) approach. This emphasizes that patterns during peaks of regional brain signals can improve traditional FC maps by dividing them into multiple transient patterns that can be observed at different intervals [12]. Such complex analysis promises deeper insights into the neural mechanisms of addictive behavior, including cocaine addiction. Moreover, as a data-driven approach, CAP analysis has the advantage of relying on minimal mathematical assumptions.

CAP analysis, which utilizes k-means clustering, generates dynamic CAPs that exhibit greater consistency than those identified using the sliding-window correlation technique, a widely used method for assessing dFC. This suggests that CAP analysis may be more effective in capturing the subtle neural changes associated with CUD, underscoring its potential for providing significant insight into the disorder [13]. These advantages have led to its increasing use in the study of abnormal network dynamics in neurological disorders [14, 15] and in studies of twins [16]. More recently, it has been used to study the reproducibility of fMRI data [17].

Although studies have examined substance use disorders (e.g., Alcohol addiction, [18]) and behavioral disorders (e.g., Internet gaming disorder[19]) and have provided evidence of abnormalities in dynamic brain configurations throughout the brain, no previous study to the best of our knowledge has leverage CAP analysis to examine brain dynamics in CUD. Therefore, it is imperative to investigate how the resting-state whole-brain coactivate over time and in different States.

This study aims to characterize the state dynamic profiles and sFC characteristics of CUD using CAP analysis. CAP analysis can capture transient brain states and their coactivation patterns, offering a detailed characterization of brain dynamics in CUD, potentially leading to an improved understanding of the disorder. We hypothesize that individuals with CUD will exhibit distinct coactivation patterns and alterations in both dynamic and sFC compared to HC, with significant differences in their temporal dynamics, reflecting their clinical characteristics and behavioral patterns.

## 2. Materials and methods

### 2.1. Dataset and participate

We used publicly available data, the Mexican dataset of cocaine use disorder patients (SUDMEX_CONN) for this study (https://openneuro.org/datasets/ds003346/versions/1.1.2). Eleven subjects were excluded on the basis that their maximum translational or rotational was greater than 2 mm or 2° as well as framewise displacement (FD) > 0.5 [20]. After excluding subjects with large head motion, 56 CUD participants and 57 HC participants remained for the subsequent analysis. The demographic information was provided in Table 1, and the statistical differences between the two groups used a two-sample t-test for age and education, and a Chi-squared test for gender.

**Table 1.** The demographic information.

| HC | CUD | P-Value |
| --- | --- | --- |

|  | 57 | 56 |  |
| --- | --- | --- | --- |
| Age [years] | 30.98 ± 8.07 | 31.77 ± 7.52 | 0.594 <sup>a)</sup> |
| Gender (M/F) | 46 / 11 | 49 / 7 | 0.323 <sup>b)</sup> |
| Education years | 12.67 ± 3.302 | 11.30 ± 3.068 | 0.025 <sup>a)</sup> * |
| years of cocaine use | - | 10.42 ± 6.90 | - |
| CUD age onset | - | 21.19 ± 5.94 | - |

Data are expressed as mean ± SD (SD: standard deviation). a) two-sample t-test; b) chi-square cross-table test.

### 2.2 Image acquisition

Imaging was conducted using a Philips Ingenia 3 T MR system (Philips Healthcare, Best, The Netherlands, and Boston, MA, USA), with a 32-channel dS Head coil. Resting-state fMRI sequences were acquired using a gradient recalled (GE) echo planar imaging (EPI) sequence with the following parameters: dummies = 5, repetition time (TR)/echo time (TE) = 2000/30.001 ms, flip angle = 75°, matrix = 80×80, field of view = 240 mm^2^, voxel size = 3×3×3 mm^3^, slice acquisition order = interleaved (ascending), number of slices = 36, phase encoding direction = AP. The total scan time of the rs-fMRI session was 10 min with a total of 300 volumes acquired. All participants were instructed to keep their eyes open, to relax while not thinking about anything in particular, and not to fall asleep. T1- weighted were acquired using a three-dimensional FFE SENSE sequence, TR/TE = 7/3.5 ms, field of view = 240 mm^2^, matrix = 240×240 mm, number of slices = 180, gap = 0, plane = sagittal, voxel = 1×1×1 mm^3^.

### 2.3 Preprocessing

The resting-state fMRI data were preprocessed using DPABI (http://rfmri.org/dpabi) following these steps: 1) Removal of the first 10 time points; 2) Slice timing correction; 3) Realignment; 4) Brain extraction; 5) Coregistration of T1 image to functional image; 6) T1 segmentation using DARTEL; 7) Normalization of the functional images by T1 DARTEL; 8) Nuisance regression, including 24 head motion parameters, mean white matter (WM) and mean cerebrospinal fluid (CSF) signal, both with and without global signal regression (GSR); 9) Application of band-pass filtering, ranging from 0.01 Hz to 0.08 Hz; 10) Smoothing with an 8 mm FWHM kernel.

### 2.4 Coactivation pattern analysis

In this study, the CAP analysis was performed using homemade scripts in MATLAB (https://ww2.mathworks.cn/products/matlab.html), with the main code available at https://github.com/davidyoung1994/CoactivationPattern. The CAP analysis is a data-driven method that utilizes K-means clustering to identify recurring whole-brain coactivation States.

In brief, there were 290 volumes for each subject’s preprocessed fMRI data, and each volume was characterized by the activation level of 400 ROIs using Yeo’s 7 network parcellation [21, 22].The time series of each ROI was first normalized using z-score independently, and the absolute value of Z indicated the activation deviation from its baseline. Then, K-means clustering was performed based on all volumes of the 56 CUD subjects and 57 HC. The cluster number K was selected from 2 to 21 with a step length of 1. The clustering algorithm was repeated 100 times with a new initial cluster centroid for each K-value, and the results with the lowest within-cluster sums of point-to-centroid distances were used. Volumes with similar co-activation profiles were grouped into the same CAP states. Frames assigned to the same CAP state were averaged and divided by the within-cluster standard deviation to generate the normalized CAP maps (Z-maps) at the group levels.

### 2.5 State temporal dynamics metrics

To evaluate the dynamic properties within and between CAP states, four dynamic metrics were calculated at the individual level: 1) Fraction of Occupied Time (FOT): This metric represents the proportion of total time that each group spends in a particular CAP state (brain state) over the whole time series; 2) Persistence (PST): This metric represents the average duration that a specific brain state remains stable for each group before transferring to another State(mean volume-to-volume maintenance of one CAP state); 3) Counts: This metric represents the number of times each group transitions into a specific brain state (how many times one State occurred during the whole scan); and 4). Transition probability matrix is the probability that one volume within State A transfers to the next volume belonging to State B, with a non-zero diagonal as the volume within State A could still stay within State A for the next volume.

### 2.6 Statistical analysis

The demographic characteristics in age and education years were tested using the two-sample T-test, and gender was tested using the chi-squared cross-table test. As for the CAP metrics, two-sample t-tests were performed to estimate between-group differences, while controlling age and gender as covariates. False discovery rate (FDR) correction (q = 0.05) was used to account for the multiple comparisons.

The relationships between CAP metrics and cocaine abuse information including age of initiation (years of cocaine use), and duration of continued use (CUD age of onset) were estimated using partial correlation with age, gender, and education years as covariates. The results are corrected here using the permutation test, which is a non-parametric statistical method that assesses the significance of differences by constructing a null hypothesis to study the distribution of sample differences.

### 2.7 Exploratory analysis

#### 2.7.1 Stationary functional connectivity

To investigate the relationship between the CAP states and the stationary functional network, we identified regions of interest (ROIs) by extracting the brain regions with the highest peak values across all six CAP states. We then calculated the functional connectivity between the selected ROIs and the whole brain to identify networks that were associated with these ROIs between the two groups. Statistical significance for within group was set to p < 0.001 and the multiple comparison correction was applied to the between-group differences (FDR q = 0.05).

#### 2.7.2 Characterization of Networks and Behavioral Analysis

We employed meta-analytic term mapping from the Neurosynth database [23] to functionally characterize and differentiate the identified regions with significant FC between CUD and HC. Peak coordinates from these regions served as seeds to generate maps of meta-analytic functional connectivity (MFC) and meta-analytic co-activation (MCoact). Then top 10 associations between the seed regions and specific cognitive, psychological, or behavioral terms, along with their corresponding correlation coefficients (r) are obtained.

## 3. Results

### 3.1. Coactivation patterns and brain States

This study aimed to explore the effects of cocaine on the dynamic activity of the human brain using CAP analysis. Six coactivation patterns were generated from 56 HC subjects and 57 CUD subjects using temporal k-means clustering (Fig. 1A). Among the six CAP states, five CAP states showed high spatial similarity between the CUD and HC groups (r ≥ 0.80). Brain regions in these States that belong to the same functional network tend to be activated or deactivated simultaneously. Specifically, our results showed that State 1 primarily demonstrates activation in the default mode network (DMN); State 2 demonstrates activation in the ventral attention network (VAN). State 3 is characterized by slight activation in the frontoparietal network (FPN) and negative activation in the somatomotor network (SMN). State 4 exhibits the opposite pattern to State 3, with activation in the somatomotor network (SMN) and visual network (VN). State 5 demonstrates activation in the dorsal attention network (DAN). Interestingly State 6 exhibited an opposite spatial pattern between HC and CUD, i.e., an activation region of CUD with exactly negative HC activation (r = −0.93). State 6 in the HC group showed mainly activation in the limbic network (LN) as well as negative activation in FPN, showing spatial similarity to State 2 in the CUD group (r = 0.60) as shown in Fig. 1B, whereas State 6 in the CUD group showed stronger activation in FPN and VN.

**Fig. 1.**
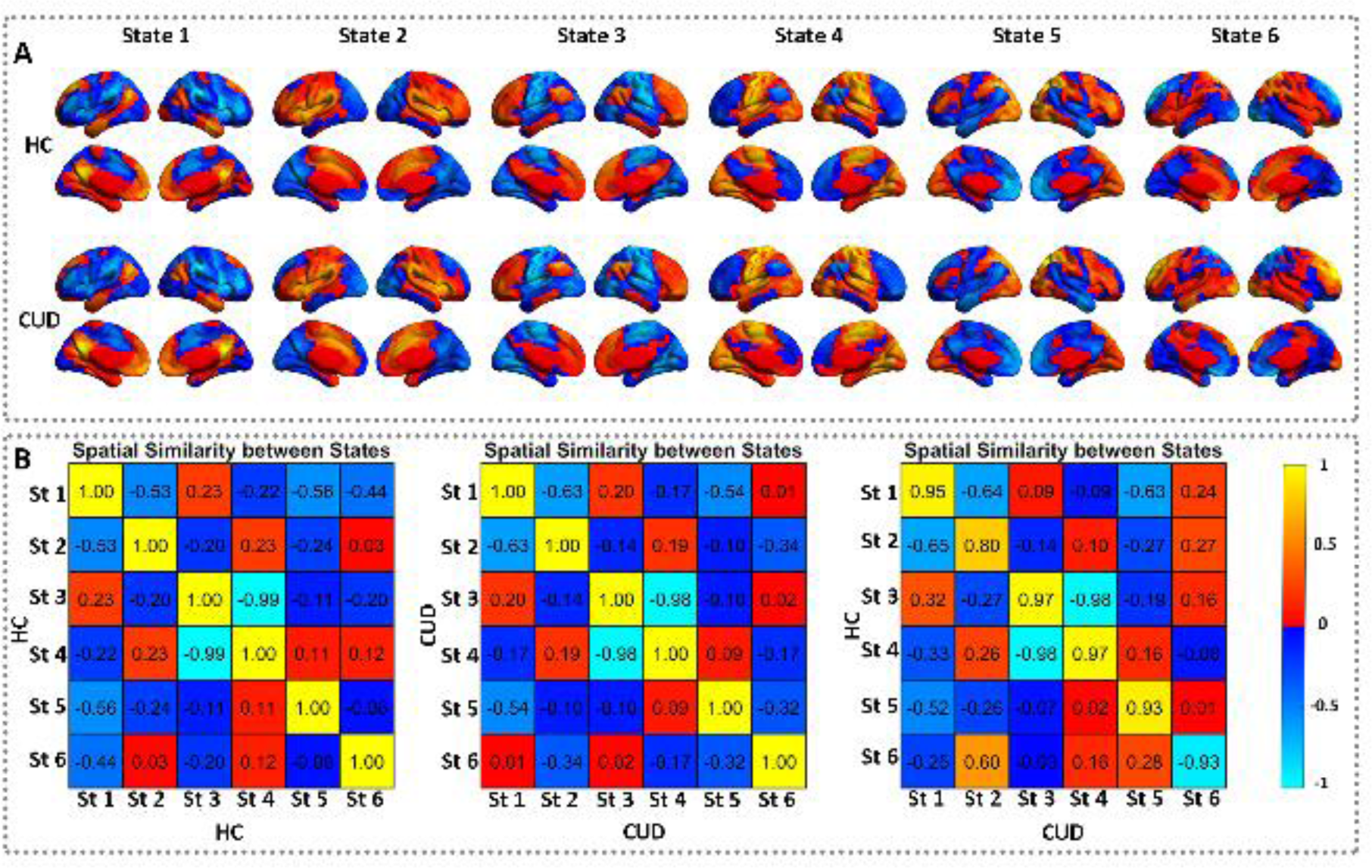
The spatial patterns for the six CAP states in cocaine addiction disease (CUD) and health control (HC) group. A) The first row shows the six states of the HC group, representing the activation of DMN, VAN, FPN, SMN& VN, DAN and LN in sequence. The second row shows the six states of the CUD group. These brain states were normalized at the group level and represents the z-statistic value. B) The CAP spatial similarity between states. The spatial similarity was measured by the Pearson correlation.

### 3.2. State temporal dynamic differences between CUD and HC

States 1 to 5 showed consistent trends in spatial CAPs; hence, we assessed the corresponding State temporal dynamics. The State temporal dynamics were compared between the CUD and HC groups using a two-sample t-test, with age, gender, and education (in years) as covariates. The results of group comparisons are presented in Fig. 2.

**Fig. 2.**
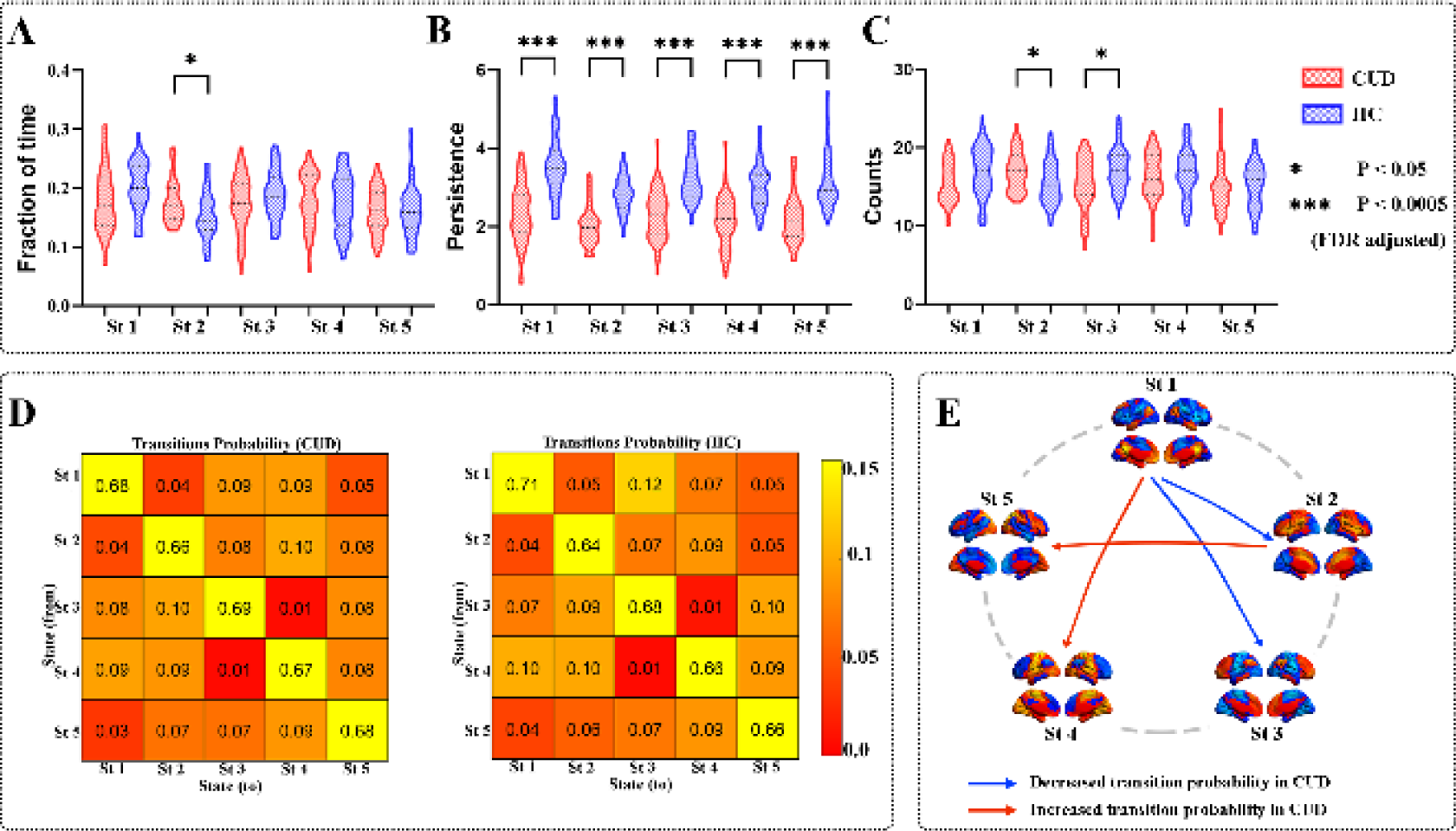
State temporal dynamic differences between CUD and HC. A-C) The group differences in fraction of time, persistence, and counts, respectively. Two-sample t-tests were performed with age, gender and education as covariates. Error-bar is the standard error. ∗ indicates p < 0.05, and ∗∗∗ indicates p < 0.0005 with FDR correction. B) The group average transition probability matrix for CUD and HC. C) The group differences in transition probability between CUD and HC. For the transition probability, the red arrow means higher transition probability in the CUD group, and the blue arrow means lower transition probability in the CUD group.

FOT: This metric represents the proportion of total time that each group spends in a particular brain state. Specifically, the CUD group showed a significantly increased FOT in State 2 (VAN), with approximately 28%, compared to the HC group’s approximately 22% (p < 0.05, FDR corrected), as shown in Fig. 2A.

PST: This metric represents the average duration that a specific brain state remains stable for each group. All five states showed significantly reduced PST in the CUD group compared to the HC group. Specifically, the CUD group exhibited an average of approximately 4 seconds, whereas the HC group showed approximately 6 seconds in States 1 (DMN) and 5 (DAN). In contrast, in State 2 (VAN), State 3 (FPN), and State 4 (SMN), the HC group showed approximately 5 seconds (p < 0.005, FDR corrected), as shown in Fig. 2B.

Counts: This metric represents the number of times each group transitions into a specific brain state. Fig. 2C shows that the CUD group transitions approximately 24 times into State 2 and 20 times into State 3, whereas the HC group transitions approximately 20 times into State 2 and 24 times into State 3 (p < 0.05, FDR corrected). Regarding the transition probabilities between CAP states, the CUD group exhibited higher values for transitions from State 1 to State 4 and from State 2 to State 5. Conversely, the CUD group showed lower probabilities for transitions from State 1 to State 2 and from State 1 to State 3 (Fig. 2D and 2E).

### 3.3 The relationships between State temporal dynamics and clinical variables

The clinical relevance with state temporal dynamics was evaluated in the CUD group, using partial correlation with age, gender and education years controlled. As shown in Fig. 3, the persistence of State 5 (r = −0.2856, p = 0.0382, corrected with permutation tests) was negatively correlated with the years of consumption, while the persistence of State 6 (r = 0.2783, p = 0.0436, corrected with permutation tests) was positively correlated with the years of consumption.

**Fig. 3.**
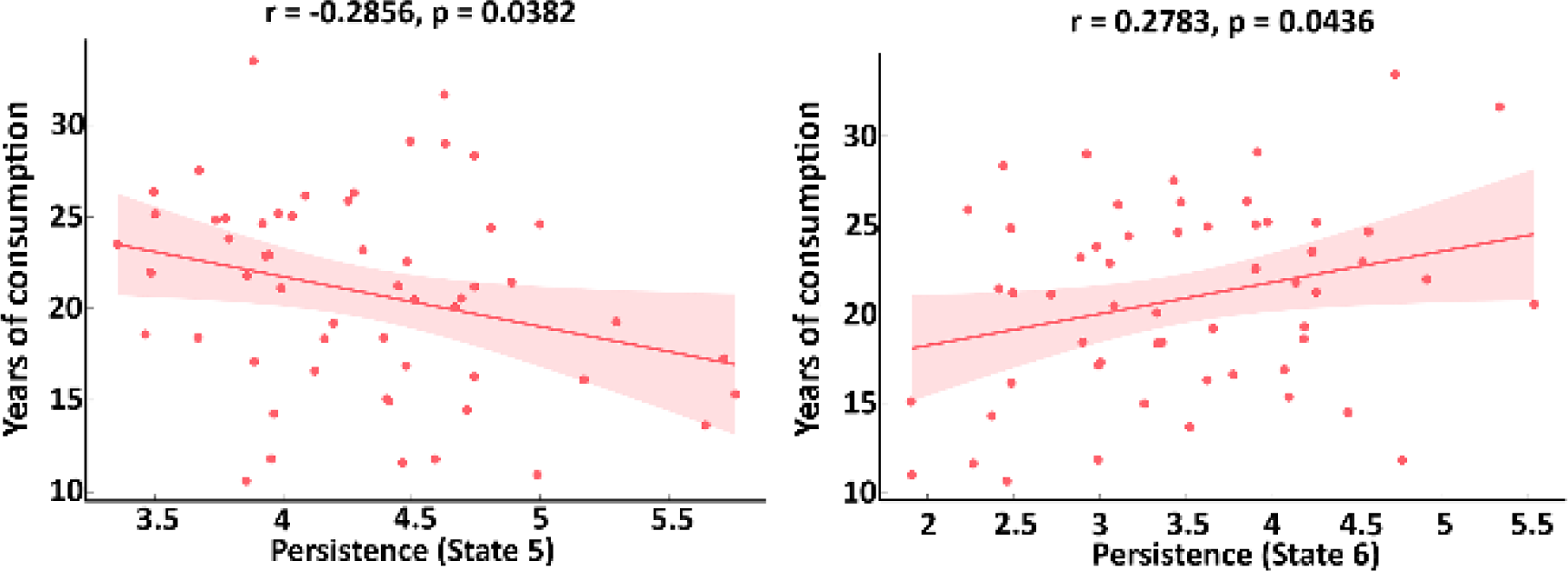
The relationships between state temporal dynamics and clinical data in CUD. Age, gender and education years were controlled as covariate. The shadow represents the 95% confidence interval.

### 3.4. Stationary functional network connectivity

Comparative FC analyses between CUD and HC states revealed both decreased and increased FC in different networks in each CAP States 1, 3, 4 and 5 (see Fig. 4A) The peak values from each state used to compute the FC are presented in Table 2. Notably, the FC patterns closely mirrored the activation patterns observed in the CAP states, with minimal deviations. Specifically, for State 1, a significantly increased region (right supramarginal gyrus: x = 48, y = −42, z = 42, t = 4.08) was identified in the DMN. State 3 exhibited a decreased region (Left Superior frontal gyrus, medial: x = 0, y = 54, z = 3, t = −3.7) was identified within the anterior DMN/ cognitive control. In State 4, a reduced region (Right precentral gyrus: x = 30, y = −15, z = 56, t = −2.9) was observed in the SMN. Finally in State 4, an increase region (Right lingual gyrus: x = 3, y = −66, z = 0, t=3.9), a region within the VSN. These results highlight the nuanced changes in activation and connectivity within different functional networks associated with CAP states when compared to the HC.

**Fig. 4.**
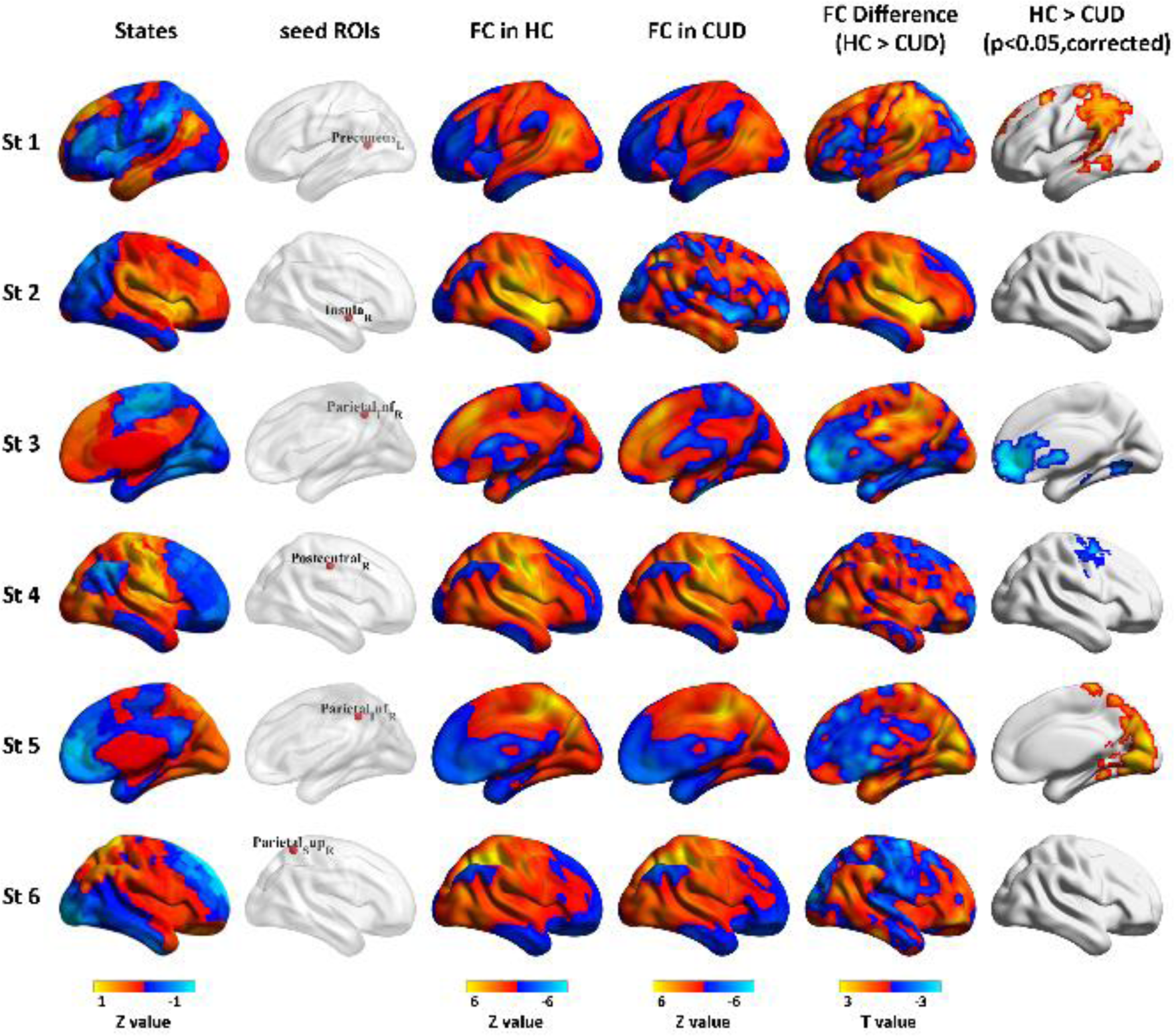
Connectivity maps based on six seeds from the Six state, these seeds (third and fourth columns) resembled the maps of dynamic brain states. Regions with significantly reduced/increased connectivity (FDR corrected for multiple comparisons at p < 0.05) are shown in the last column.

**Table 2.**
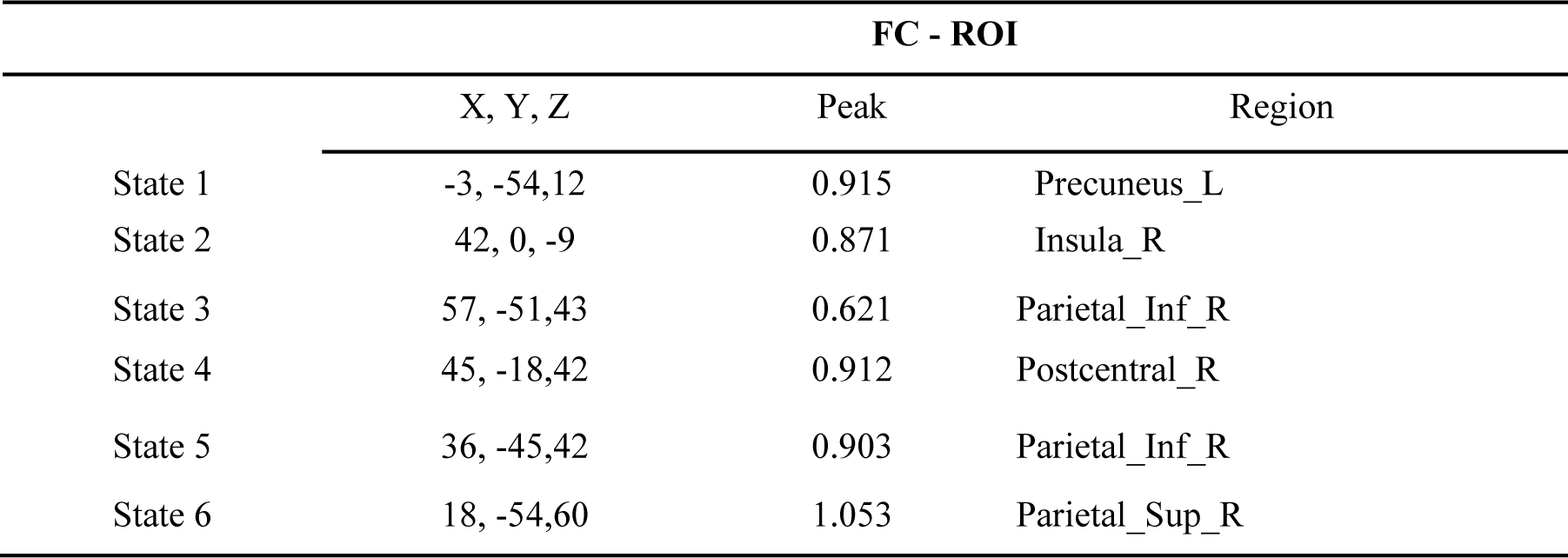
The highest peak activation for the spatial maps of the six States.

### 3.5 Functional network characterization of the identified regions

We explored meta-analytic functional connectivity (MFC) and meta-analytic co-activation (MCoact) and their functional/cognitive/behavioral associations with the four significant brain regions across different states (Fig. 5). In the DMN, the right Supramarginal Gyrus in State 1 showed the strongest connection with “working memory” and “calculation” (MFC r = 0.4, MCoact r = 0.55), suggesting their dominance in this state (Fig. 5A). Similarly, the left Superior Frontal Gyrus within the anterior DMN during State 3 exhibited the highest MFC (r = 0.39) and MCoact (r = 0.39) with “autobiographical memory” and “self-referential” processing, highlighting their importance (Fig. 5B). The right Precentral Gyrus within the SMN displayed the strongest associations with “movements” and “motor imagery” in both MFC (r = 0.58) and MCoact (r = 0.62), underlining its role in motor function in State 4 (Fig. 5C). Finally, the right Lingual Gyrus in the VSN during State 5 showed the highest MFC with vision and eye movements (r = 0.29), but weaker MCoact (Fig. 5D), compared to other regions, suggesting a general association with visual processing in this state.

**Fig. 5.**
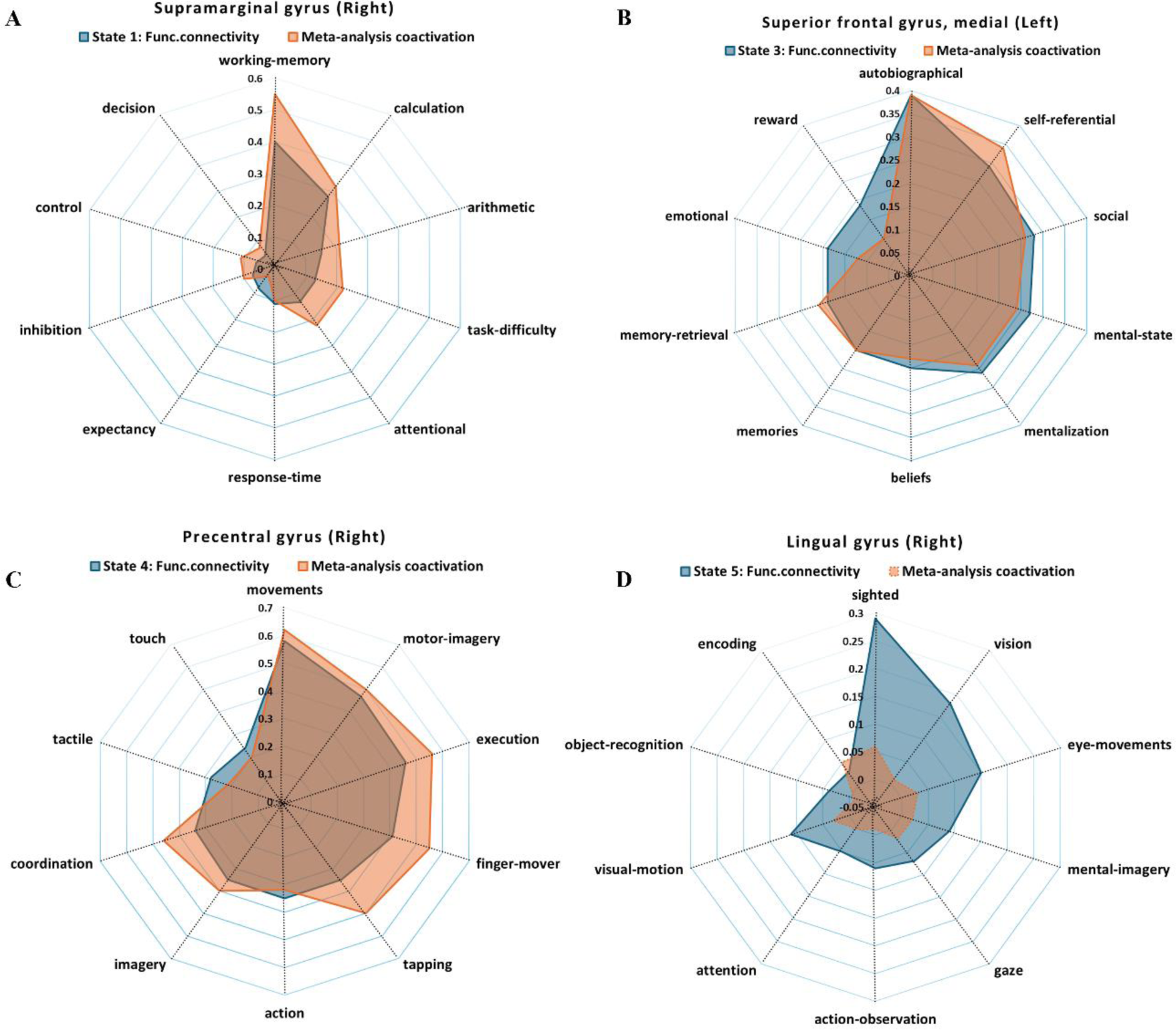
Spider plots Functional characterization of the functionally significant regions obtained in specific brain states with meta-analysis coactivation across various cognitive functions in different brain regions. (A)-D indicates different CAPS brain States with specific functional network characterization term associations obtained from Neurosynth database. Each figure shows the top ten coefficients associated with the brain seed regions.

## 4. Discussion

This study aims to characterize the State dynamic profiles and sFC characteristics of CUD using CAP. By uncovering subtle changes in brain activation and connectivity across different functional networks and states, it seeks to deepen the understanding of neural mechanisms and brain networks that underlie resting-state patterns in individuals diagnosed with CUD. We found spatial similarities in five of six different CAP states between CUD and HC groups, with a notable exception in State 6. In addition, the temporal dynamics of CUD was significantly lower compared to HC exhibited significant differences in all five persistence brain States. Interestingly, States 5 and 6 of the CAPs are significantly associated with years of consumption. We also observed significant changes in sFC in regions such as the right supramarginal gyrus, left superior frontal gyrus, right precentral gyrus, and right lingual gyrus, each associated with distinct cognitive and behavioral processes.

### 4.1 Dynamic Coactivation Patterns

We identified six CAP patterns, five of which demonstrated high spatial similarity between CUD and HC. The ability of CAPs to capture the transient States of the brain was evident as regions within the same functional network activated or deactivated simultaneously. A notable finding emerged, revealing an entirely opposite spatial pattern in State 6 between HC and CUD, suggesting a potential negative correlation between the two groups in terms of functional network activation. In the HC group, State 6 was characterized by activation in the DAN and LN, in conjunction with deactivation in the FPN. In contrast, the CUD group showed stronger activation in the FPN and the VN in State 6.

The spatial similarity matrix between the HC and CUD groups indicated a high degree of spatial correspondence (r = 0.60) between State 6 in the HC subjects and State 2 in the CUD subjects. This suggests that active states in CUD individuals, closely resembling State 6 in HC subjects, may have converged to State 2 in CUD subjects.

Given the association between the FPN and higher cognitive functions, this suggests that cocaine-induced neuronal excitation in CUD patients resulted in increased activation of the FPN during the resting state. This hypothesis is supported by the observation of greater activation of the FPN in State 6 in CUD subjects compared to HC subjects, consistent with our previous meta-analysis[24]. Moreover, this state distinction could imply a neuroadaptive response to chronic cocaine exposure, which may relate to the compulsive behavior observed in CUD patients [1].

### 4.2 Temporal Dynamics: Implications for Brain Function in CUD

The primary results from state temporal dynamics between individuals with CUD and HC provided crucial insights into the potential neural mechanisms underlying CUD. The findings reveal significant alterations in the temporal dynamics of brain states in the CUD group, highlighting the neural correlates of cognitive and behavioral dysfunctions associated with cocaine addiction.

The high FOT metric engaged within the VAN in CUD suggests heightened attentional demands and may reflect the persistent focus on drug-related cues, which is characteristic of addiction [25]. Previous studies have shown that the VAN is crucial for detecting salient stimuli and reallocating attentional resources [26]. The prolonged engagement of the VAN in CUD could indicate an adaptive response to the constant need to process and respond to drug-related stimuli, potentially at the expense of other cognitive functions.

The analysis reveals that all five brain States exhibit significantly reduced PST in the CUD group compared to the HC group. Specifically, the CUD group shows an average persistence of approximately 4 seconds in States 1 (DMN) and 5 (DAN), whereas the HC group shows approximately 6 seconds in these states. For States 2 (VAN), 3 (FPN), and 4 (SMN), the HC group demonstrates an average persistence of around 5 seconds, compared to 4 seconds in the CUD group. The DMN as well as the other higher-order networks are implicated in self-referential thinking and mind-wandering, cognitive controls and inhibition control and its disruption may contribute to the impaired introspection and self-awareness observed in CUD [7, 27].

The increased transitions into the VAN and decreased transitions into the FPN in CUD suggest a shift in the balance of attention and executive control networks. In addition, the frequent transitions into the VAN further emphasize the hyper-responsiveness to salient, often drug-related, stimuli. Also, the CUD group exhibits higher probabilities for transitions from State 1 (DMN) to State 4 (SMN) and from State 2 (VAN) to State 5 (DAN). Conversely, the CUD group shows lower probabilities for transitions from State 1 to State 2 and from State 1 to State 3 (FPN). These findings suggest a disruption in the typical flow of neural activity between major brain networks in CUD.

Moreover, the increased transitions from the DMN to the SMN may reflect a heightened readiness for motor responses, potentially related to drug-seeking. Lower transition probabilities between the DMN and attentional/executive networks (VAN and FPN) may indicate impaired switching between introspective and externally focused states, contributing to the cognitive inflexibility observed in CUD.

### 4.3 Clinical Correlation and Implications

The observed negative correlation between the PST of State 5 and years of consumption suggests a link between prolonged cocaine use and changes in brain networks. This may reflect dysfunction of the DMN and DAN as a result of long-term cocaine use and is consistent with existing literature highlighting disruptions in these networks associated with substance use disorders [28, 29]. Conversely, the positive correlation between State 6 persistence and years of consumption suggests a potential adaptive or compensatory response in certain brain networks.

Also, the PST of State 6, which correlated with of cocaine use, suggests that specific neural adaptations may be occurred to cope with chronic exposure to the drug. This adaptive response could involve changes in network connectivity, or other mechanisms aimed at maintaining cognitive and functional abilities despite the persistent effects of cocaine on the brain.

### 4.4 Stationary FC and functional network characterization

The relationship between CAP states, sFC and their association with behavioral/cognitive/functional terms underscores the proposition that changes in these brain States may be closely associated with persistence alterations in the functional network connectivity profiles. In the right supramarginal gyrus of State 1 FC with meta-analysis coactivation reveals that “*working memory*” and “*calculation*” show higher connectivity in individuals with CUD. This association suggests that these cognitive functions are more engaged or dysregulated in this population. Previous studies have demonstrated that working memory deficits and impaired higher executive functions are implicated cognitive impairments in cocaine addiction, often linked to the dysfunction higher-order networks [25, 30]. The high association in the region may reflect a compensatory mechanism or heightened cognitive demands placed on these functions in individuals with CUD.

In State 3, the medial left superior frontal gyrus exhibited strong association with “*autobiographical memory*” and “*self-referential*” processing in individuals with CUD. These findings suggest an increased engagement in self-related processing, which may be linked to the heightened focus on drug-related cues and cravings typical in CUD especially in the DMN. The medial prefrontal cortex, including the superior frontal gyrus, is critically involved in self-referential processing and is often hyperactive in substance use disorders [28]. In addition, the superior frontal gyrus was shown to be associated with cravings in response to the cocaine cues [31]. Neurochemical imaging studies have also shown that drug addicts exhibit a marked decrease in dopamine D2 receptors and dopamine release, which is associated with reduced regional activity in key brain regions, including the dorsolateral prefrontal cortex involved in executive functions [32].

In State 4, the right precentral gyrus reveals a higher with “*movements*” and “*motor imagery*”. This region is a key component of the motor network and may reflect enhanced motor planning and execution associated with drug-seeking behavior. Studies have shown that motor brain regions in drug abusers are often more active during tasks involving drug-related stimuli, indicating a strong link between motor planning and drug use [32]. In addition, pronounced changes in bilateral sensorimotor control are associated with chronic cocaine abuse, resulting in functional changes in all movement-related brain networks [33]. This also includes negative effects on fine motor skills, as measured by bimanual tasks or maneuvers that require simultaneous coordination of limbs [34].

Results from the right lingual gyrus indicate of State 5 associated highly with “*vision*” and “*visual motion*” CUD individuals. This association suggests enhanced processing of visual information relating to the salience of drug-related visual cues. The lingual gyrus is part of the visual processing network and is associated with the recognition and processing of visual stimuli. High association in this region may reflect the heightened attention and sensitivity to drug-related cues observed in CUD, contributing to strong visual triggers for craving.

The functional significance of drug cue-induced occipital cortex activation remains unclear. Some posit a role in reward processing, while others emphasize attentional mechanisms. Regardless, research consistently demonstrates robust cue discrimination by the visual cortex in individuals with substance dependence [35–37].

### 4.5 Limitations and Future Directions

Firstly, due to the cross-sectional nature of our study, it is only possible to establish causal relationships to a limited extent. The dynamic changes observed in the CAP states could be a consequence of prolonged cocaine use, but it remains unclear whether these changes are preexisting and predispose individuals to CUD. Longitudinal studies are needed to reveal the temporal sequence of these brain changes and their impact on the development and progression of addiction.

Furthermore, the demographic homogeneity of our study may limit the generalizability of our results. The inclusion of different populations, taking into account factors such as polydrug use and psychiatric comorbidities, would improve the external validity of our results and provide a more comprehensive picture of the neural dynamics associated with CUD. Despite these limitations, our study represents a significant advance in examining the dynamic aspects of brain function in individuals with CUD.

## 5. Conclusion

This study highlights the importance of dynamic CAP analysis in understanding CUD. By examining resting-state brain activity, significant alterations in brain networks and their temporal dynamics were identified in individuals with CUD compared to healthy controls. The findings revealed heightened attentional demands and disrupted transitions between cognitive states coupled with altered, particularly affecting the DMN, VAN and FPN. These investigation into the neural mechanisms of CUD highlight the potential for CAP analysis to inform more helpful approaches in studying addiction.

## Ethics approval and consent to participate

Not applicable

## Consent for publication

Not applicable

## Availability of data and materials

The dataset supporting the conclusions of this article is available in the Openneuro repository (https://openneuro.org/datasets/ds003346/versions/1.1.2.). The main code about CAP analysis (https://github.com/davidyoung1994/CoactivationPattern.)

## Authors’ contributions

BK-B conceptualized and interpreted results and drafted the manuscript. YX carried out the main computations and analyses, and interpreted results. HY provided scripts for the main CAPs analysis. PW and BB edited and supervised the manuscript. All authors contributed to the manuscript for important intellectual content, approved the final manuscript.

## Data Availability

All data produced are available online at the Openneuro repository

https://openneuro.org/datasets/ds003346/versions/1.1.2.

## Acknowledgements

Not applicable

## References

1. Volkow ND, Michaelides M, Baler R. The neuroscience of drug reward and addiction. Physiol Rev. 2019;99.

2. Ma L, Steinberg JL, Cunningham KA, Lane SD, Bjork JM, Neelakantan H, et al. Inhibitory behavioral control: A stochastic dynamic causal modeling study comparing cocaine dependent subjects and controls. Neuroimage Clin. 2015;7:837–47.

3. Kelly C, Zuo XN, Gotimer K, Cox CL, Lynch L, Brock D, et al. Reduced interhemispheric resting state functional connectivity in cocaine addiction. Biol Psychiatry. 2011;69:684–92.

4. Biswal B, Zerrin Yetkin F, Haughton VM, Hyde JS. Functional connectivity in the motor cortex of resting human brain using echo-planar mri. Magn Reson Med. 1995;34.

5. Fox MD, Raichle ME. Spontaneous fluctuations in brain activity observed with functional magnetic resonance imaging. Nature Reviews Neuroscience. 2007;8.

6. Greicius MD, Supekar K, Menon V, Dougherty RF. Resting-state functional connectivity reflects structural connectivity in the default mode network. Cerebral Cortex. 2009;19.

7. Geng X, Hu Y, Gu H, Salmeron BJ, Adinoff B, Stein EA, et al. Salience and default mode network dysregulation in chronic cocaine users predict treatment outcome. Brain. 2017;140:1513–24.

8. Klugah-Brown B, Di X, Zweerings J, Mathiak K, Becker B, Biswal B. Common and separable neural alterations in substance use disorders: A coordinate-based meta-analyses of functional neuroimaging studies in humans. Hum Brain Mapp. 2020. 10.1002/hbm.25085.

9. Klugah-Brown B, Jiang C, Agoalikum E, Zhou X, Zou L, Yu Q, et al. Common abnormality of gray matter integrity in substance use disorder and obsessive-compulsive disorder: A comparative voxel-based meta-analysis. Hum Brain Mapp. 2021;42.

10. Honey CJ, Sporns O, Cammoun L, Gigandet X, Thiran JP, Meuli R, et al. Predicting human resting-state functional connectivity from structural connectivity. Proc Natl Acad Sci U S A. 2009;106.

11. Zalesky A, Fornito A, Cocchi L, Gollo LL, Breakspear M. Time-resolved resting-state brain networks. Proc Natl Acad Sci U S A. 2014;111.

12. Liu X, Duyn JH. Time-varying functional network information extracted from brief instances of spontaneous brain activity. Proc Natl Acad Sci U S A. 2013;110.

13. Hutchison RM, Womelsdorf T, Allen EA, Bandettini PA, Calhoun VD, Corbetta M, et al. Dynamic functional connectivity: Promise, issues, and interpretations. Neuroimage. 2013;80:360– 78.

14. Kaiser RH, Kang MS, Lew Y, Van Der Feen J, Aguirre B, Clegg R, et al. Abnormal frontoinsular-default network dynamics in adolescent depression and rumination: a preliminary resting-state co-activation pattern analysis. Neuropsychopharmacology. 2019;44.

15. Ma X, Zhuo Z, Wei L, Ma Z, Li Z, Li H. Altered Temporal Organization of Brief Spontaneous Brain Activities in Patients with Alzheimer’s Disease. Neuroscience. 2020;425.

16. Yao X, Klugah-Brown B, Yang H, Biswal B. Structural and functional network analysis of twins using fMRI data. Cerebral Cortex. 2023. 10.1093/cercor/bhad345.

17. Yang H, Zhang H, Di X, Wang S, Meng C, Tian L, et al. Reproducible coactivation patterns of functional brain networks reveal the aberrant dynamic state transition in schizophrenia. Neuroimage. 2021;237.

18. Wang KS, Brown K, Frederick BB, Moran L V., Olson D, Pizzagalli DA, et al. Nicotine acutely alters temporal properties of resting brain states. Drug Alcohol Depend. 2021;226.

19. Niu X, Gao X, Zhang M, Dang J, Sun J, Lang Y, et al. Static and dynamic changes of intrinsic brain local connectivity in internet gaming disorder. BMC Psychiatry. 2023;23.

20. Power JD, Barnes KA, Snyder AZ, Schlaggar BL, Petersen SE. Spurious but systematic correlations in functional connectivity MRI networks arise from subject motion. Neuroimage. 2012;59:2142–54.

21. Schaefer A, Kong R, Gordon EM, Laumann TO, Zuo XN, Holmes AJ, et al. Local-global parcellation of the human cerebral cortex from intrinsic functional connectivity mri. Cerebral Cortex. 2018;28.

22. Thomas Yeo BT, Krienen FM, Sepulcre J, Sabuncu MR, Lashkari D, Hollinshead M, et al. The organization of the human cerebral cortex estimated by intrinsic functional connectivity. J Neurophysiol. 2011;106:1125–65.

23. Yarkoni T, Poldrack RA, Nichols TE, Van Essen DC, Wager TD. Large-scale automated synthesis of human functional neuroimaging data. Nat Methods. 2011;8.

24. Taebi A, Becker B, Klugah-Brown B, Roecher E, Biswal B, Zweerings J, et al. Shared network-level functional alterations across substance use disorders: A multi-level kernel density meta-analysis of resting-state functional connectivity studies. Addiction Biology. 2022;27:1–10.

25. Goldstein RZ, Volkow ND. Dysfunction of the prefrontal cortex in addiction: Neuroimaging findings and clinical implications. Nat Rev Neurosci. 2011;12:652–69.

26. Vossel S, Geng JJ, Fink GR. Dorsal and ventral attention systems: Distinct neural circuits but collaborative roles. Neuroscientist. 2014;20.

27. Buckner RL, Andrews-Hanna JR, Schacter DL. The brain’s default network: Anatomy, function, and relevance to disease. Annals of the New York Academy of Sciences. 2008.

28. Sutherland MT, McHugh MJ, Pariyadath V, Stein EA. Resting state functional connectivity in addiction: Lessons learned and a road ahead. NeuroImage. 2012.

29. Zhang R, Volkow ND. Brain default-mode network dysfunction in addiction. NeuroImage. 2019;200.

30. Ersche KD, Barnes A, Simon Jones P, Morein-Zamir S, Robbins TW, Bullmore ET. Abnormal structure of frontostriatal brain systems is associated with aspects of impulsivity and compulsivity in cocaine dependence. Brain. 2011;134.

31. Ernst M, Ph D, London ED, Ph D. Neural Systems and Cue-Induced Cocaine Craving. 1999.

32. Volkow ND, Fowler JS, Wang GJ, Baler R, Telang F. Imaging dopamine’s role in drug abuse and addiction. Neuropharmacology. 2009;56 SUPPL. 1.

33. Hanlon CA, Wesley MJ, Roth AJ, Miller MD, Porrino LJ. Loss of laterality in chronic cocaine users: An fMRI investigation of sensorimotor control. Psychiatry Res Neuroimaging. 2010;181:15–23.

34. Roizenblatt M, Fidalgo TM, Polizelli M, Cruz NFS da, Roizenblatt A, Jiramongkolchai K, et al. Effect of chronic cocaine use on fine motor coordination tested during ophthalmic vitreoretinal simulated performance. J Psychiatr Res. 2021;132.

35. Hanlon CA, Dowdle LT, Naselaris T, Canterberry M, Cortese BM. Visual cortex activation to drug cues: A meta-analysis of functional neuroimaging papers in addiction and substance abuse literature. Drug Alcohol Depend. 2014;143.

36. Chase HW, Eickhoff SB, Laird AR, Hogarth L. The neural basis of drug stimulus processing and craving: An activation likelihood estimation meta-analysis. Biol Psychiatry. 2011;70:785–93.

37. Schacht JP, Anton RF, Myrick H. Functional neuroimaging studies of alcohol cue reactivity: A quantitative meta-analysis and systematic review. Addiction Biology. 2013;18:121–33.

